# Benefit Expectations and Neuroprotective Prescribing for Acute Ischemic Stroke in China

**DOI:** 10.64898/2026.10.29.26366879

**Authors:** Zefeng Tan, Liping Liu, Anxin Wang, Hongyu Zhou, Meng Gao, Quan Zhou, Min Hu, Xia Meng, Anding Xu

## Abstract

**Background:** Neuroprotective therapies are frequently used for acute ischemic stroke in China. We examined associations of neurologists’ self-rated familiarity, perceived importance, and benefit expectations with reported prescribing frequency.

**Methods:** A hospital-based quota survey recruited attending-level or senior neurologists involved in acute ischemic stroke care across mainland China from May 27 to July 16, 2025. High-frequency use was defined as prescribing neuroprotective therapy to at least 60% of patients. High familiarity and importance were ratings of 4 or 5 on 5-level scales; consistently high benefit expectations required ratings of 4 or 5 in all 4 scenarios. Modified Poisson generalized estimating equations estimated adjusted prevalence ratios (PRs) with hospital-clustered robust standard errors.

**Results:** Among 1982 neurologists from 251 hospitals, 1007 (50.8%) were men and 1745 (88.0%) practiced in tertiary hospitals. High-frequency use was reported by 1596 respondents (80.5%), high familiarity by 1006 (50.8%), and consistently high benefit expectations by 1290 (65.1%). Lower familiarity with high-frequency use was reported by 743 respondents (37.5%). High familiarity (adjusted PR, 1.053; 95% CI, 1.007–1.102), high perceived importance (adjusted PR, 1.294; 95% CI, 1.148–1.458), and consistently high benefit expectations (adjusted PR, 1.198; 95% CI, 1.131–1.269) were associated with high-frequency use after adjustment for demographic and professional characteristics.

**Conclusions:** Frequent self-reported neuroprotective prescribing coexisted with lower self-rated familiarity in a substantial subgroup. Familiarity, perceived importance, and benefit expectations were associated with prescribing frequency. These cross-sectional, class-level measures cannot establish prescribing appropriateness or causality. Studies linking drug-specific observed prescribing to indications and patient outcomes are needed.

## Introduction

Neuroprotection for acute ischemic stroke (AIS) remains a persistent translational challenge. Experimental work has identified multiple potential targets, but many clinically promising interventions have not produced consistent benefit in confirmatory trials.^1,2^ Reperfusion therapy has transformed acute stroke care, yet many patients remain ineligible for reperfusion or have residual disability despite recanalization.^3,4^ These gaps sustain interest in cytoprotective strategies used either with reperfusion or when reperfusion cannot be performed. The evidence and practice environment in China is heterogeneous. Chinese guidance includes agent-specific recommendations, and recent randomized clinical trials have reported benefits for selected agents or formulations in defined populations and treatment windows.^5-8^ These findings do not establish a uniform class effect for all therapies described as neuroprotective. Consequently, a clinician’s reported use may reflect different drugs, indications, local pathways, formulary arrangements, and interpretations of an evolving evidence base. Clinical practice is influenced not only by knowledge of evidence but also by expected benefit, institutional routines, perceived barriers, and implementation context.^9^ However, the relationship between self-rated familiarity, benefit expectations, and neuroprotective prescribing among neurologists in China has not been well characterized. We conducted a nationwide cross-sectional survey to describe these constructs and prescribing patterns and to estimate their associations with high-frequency use while accounting for clustering of physicians within hospitals.

## Methods

### Data and Materials Availability

The questionnaire versions and aggregate results are included in the Supplemental Material. Deidentified participant-level data, the study protocol, and analysis code are available from the corresponding author upon reasonable request.

### Study Design and Participants

We conducted a nationwide, cross-sectional, web-based survey of neurologists in mainland China from May 27 to July 16, 2025. The hospital-based quota recruitment strategy was stratified to achieve coverage across geographic regions, hospital levels, and professional titles. The protocol targeted 1984 neurologists from 251 hospitals. Respondents in the final analytic data set represented 251 hospitals in all 31 province-level regions of mainland China. Eligible participants were attending-level neurologists or above who were directly involved in AIS diagnosis and management and had independently managed at least 10 patients with AIS during the preceding 12 months. Individuals were not eligible if they practiced outside neurology, had no routine responsibility for AIS care, had managed fewer than 10 patients with AIS during the preceding 12 months, or declined to participate in the survey. The Chinese Stroke Association coordinated survey distribution. Participation was voluntary. The final analytic sample included 1982 neurologists from 251 hospitals. The report was prepared with reference to the Strengthening the Reporting of Observational Studies in Epidemiology (STROBE) statement and applicable American Association for Public Opinion Research recommendations.^10,11^ The completed checklist is included in the Supplemental Material. The recruitment target determined the study size. Individual invitation and nonresponse dispositions were unavailable, so a participation rate could not be calculated.

### Questionnaire Development

The questionnaire was developed based on a comprehensive literature review. Expert content review was conducted to ensure the relevance and accuracy of all items. Pilot testing was then carried out with 30 physicians to evaluate the clarity and comprehensibility of the instrument. The questionnaire was developed in Chinese. An English translation was subsequently prepared for publication in the Supplemental Material. Translation discrepancies were resolved by bilingual members of the research team to ensure conceptual consistency with the original Chinese instrument. The complete Chinese questionnaire and English translation are provided in the Supplemental Material. Participants who completed the survey were eligible to receive RMB 200 in compensation, as specified in the approved consent form.

### Measures and Outcomes

The primary outcome was self-reported neuroprotective prescribing frequency in AIS, measured by Q14 on a 5-level ordered scale. In the analytic dataset, responses to Q14 were reverse-coded from the visual order in the original survey instrument to ensure higher numerical values consistently represented higher prescribing frequencies (i.e., 5 = routine use in >90% of patients, 4 = high-frequency use in 60% to 90%, 3 = selective use in 30% to 60%, 2 = low-frequency use in <30%, and 1 = almost never). High-frequency use was defined as Q14 ratings of 4 or 5, corresponding to the original questionnaire categories of high-frequency use in 60% to 90% of patients and routine use in more than 90% of patients. Thus, the primary binary outcome represented reported neuroprotective use in at least 60% of patients with AIS. Because the 60% threshold was an operational definition based on the questionnaire categories rather than an externally validated clinical cut point, alternative thresholds were evaluated in sensitivity analyses. The 30%–60% and 60%–90% response categories overlap at 60%; analyses used the selected category without reassigning responses.

Self-rated familiarity (Q10) and perceived importance (Q11) were recorded on 5-level ordered scales. For categorical analyses, high familiarity and high importance were defined as ratings of 4 or 5; ratings of 1 to 3 formed the comparison groups. Q10 assessed subjective familiarity rather than objective evidence knowledge.

Q12 contained 8 Likert items concerning attitudes and behavioral intentions. Each item was summarized using category-specific counts and percentages; the items were not combined into a continuous total score.

Expected clinical benefit was assessed separately for 4 scenarios (Q15-Q18), defined by stroke severity and receipt of reperfusion therapy. Each item used a 5-level ordered response from no benefit to marked benefit. A high expected-benefit rating was defined as 4 or 5. The primary benefit-expectation variable was consistently high expected benefit, defined as a high rating for all 4 scenarios vs any other response pattern. Scenario-specific variables were evaluated in sensitivity analyses. A conflict between the Q18 severity descriptor and its textual label was noted in the questionnaire documentation. Analyses retained its textual label; an exploratory analysis omitted Q18 from the combined benefit definition (Table S1).

Adjustment variables in the primary model were age (<40, 40-49, or ≥50 years), sex, professional title, hospital level (tertiary vs secondary), and geographic region (eastern, central, western, or northeastern China). Hospital type was not included because Q7 permitted multiple responses and could not be interpreted as a single mutually exclusive characteristic.

### Statistical Analysis

Categorical and ordinal variables were summarized as counts and percentages. Likert responses were not summarized as continuous composite scores in the primary analysis. Spearman rank correlations were used for descriptive associations between ordinal items. Confidence intervals for selected correlations were estimated using 5000 hospital-cluster bootstrap replicates. Because high-frequency use was common, the primary analysis used modified Poisson regression to estimate prevalence ratios (PRs) and 95% CIs directly rather than odds ratios.^12^ A generalized estimating equation model with a log link, Poisson variance, exchangeable working correlation structure, hospital as the clustering unit, and robust sandwich standard errors accounted for correlation among neurologists within hospitals.^13^ The model included high self-rated familiarity, high perceived importance, consistently high expected benefit, age group, sex, professional title, hospital level, and geographic region. Estimates were interpreted as cross-sectional associations.

To assess the robustness of the 60% threshold, sensitivity analyses redefined the outcome using a more stringent threshold (Q14 = 5, corresponding to routine use in >90% of patients) and a more inclusive threshold (Q14 ≥3, corresponding to use in ≥30% of patients); replaced the consistently high benefit variable with each scenario-specific benefit item; and used an independent working correlation structure. Secondary and sensitivity analyses were interpreted as exploratory.

Additional exploratory checks omitted Q18 from the combined benefit definition and excluded all 10 records flagged by repeated telephone entries (Table S1). Repeated entries alone did not establish duplicate respondents. All variables included in the primary modified Poisson GEE model had 0 missing values, so all 1982 respondents were included in the adjusted analysis. No survey weights were applied to the quota sample. Analyses were performed in Python 3.13.5 using statsmodels 0.14.6; Spearman correlations and bootstrap calculations used SciPy 1.17.0 and custom resampling code. All tests were 2-sided, with P < .05 used as a descriptive threshold; interpretation focused on effect estimates and 95% CIs.

### Ethics

The Institutional Ethics Committee of Capital Medical University Beijing Tiantan Hospital approved the study (KY2025-135-02), with local approval obtained when required. All participants provided electronic informed consent. No patient-level data were collected.

## Results

### Respondent Characteristics

The analytic file contained 1982 neurologists from 251 hospitals. Of these respondents, 1007 (50.8%) were men, 1717 (86.6%) were aged 30 to 49 years, and 991 (50.0%) were attending neurologists. A total of 1745 (88.0%) practiced in tertiary hospitals. Respondents were from eastern (643 [32.4%]), central (385 [19.4%]), western (762 [38.4%]), and northeastern (192 [9.7%]) China (Table 1).

**Table 1.** Characteristics of Respondents (N = 1982)

| Characteristic | Category | No. (%) |
|---|---|---|
| Total respondents |  | 1982 (100.0) |
| Age, y | <30 | 45 (2.3) |
|  | 30-39 | 946 (47.7) |
|  | 40-49 | 771 (38.9) |
|  | 50-59 | 201 (10.1) |
|  | ≥60 | 19 (1.0) |
| Sex | Male | 1007 (50.8) |
|  | Female | 975 (49.2) |
| Professional title | Attending neurologist | 991 (50.0) |
|  | Associate chief neurologist | 649 (32.7) |
|  | Chief neurologist | 342 (17.3) |
| Highest educational degree | Diploma/associate | 10 (0.5) |
|  | Bachelor's | 889 (44.9) |
|  | Master's | 831 (41.9) |
|  | Doctoral | 252 (12.7) |
| Total years in clinical practice | 1-5 | 171 (8.6) |
|  | 6-10 | 470 (23.7) |
|  | 11-15 | 528 (26.6) |
|  | 16-20 | 340 (17.2) |
|  | >20 | 473 (23.9) |
| Years in neurology/stroke practice | 3-5 | 288 (14.5) |
|  | 6-10 | 578 (29.2) |
|  | 11-15 | 474 (23.9) |
|  | >15 | 642 (32.4) |
| Hospital level | Class III Grade A | 1390 (70.1) |
|  | Other tertiary | 355 (17.9) |
|  | Class II Grade A | 195 (9.8) |
|  | Other secondary | 42 (2.1) |
| Geographic region | Eastern China | 643 (32.4) |
|  | Central China | 385 (19.4) |
|  | Western China | 762 (38.4) |
|  | Northeastern China | 192 (9.7) |

Note. Percentages may not sum to 100 because of rounding. Hospital type was omitted because the source item permitted multiple selections. Geographic regions followed the study's administrative classification.

### Familiarity, Benefit Expectations, and Reported Use

High self-rated familiarity was reported by 1006 respondents (50.8%), and high perceived importance was reported by 1781 (89.9%). Across the 8 Q12 attitude and behavioral-intention items, strongly agree responses ranged from 66.8% to 73.3% and agree responses from 25.7% to 31.4%; neutral or disagreement responses were uncommon. Full 5-category distributions are reported in Table 2.

**Table 2.**
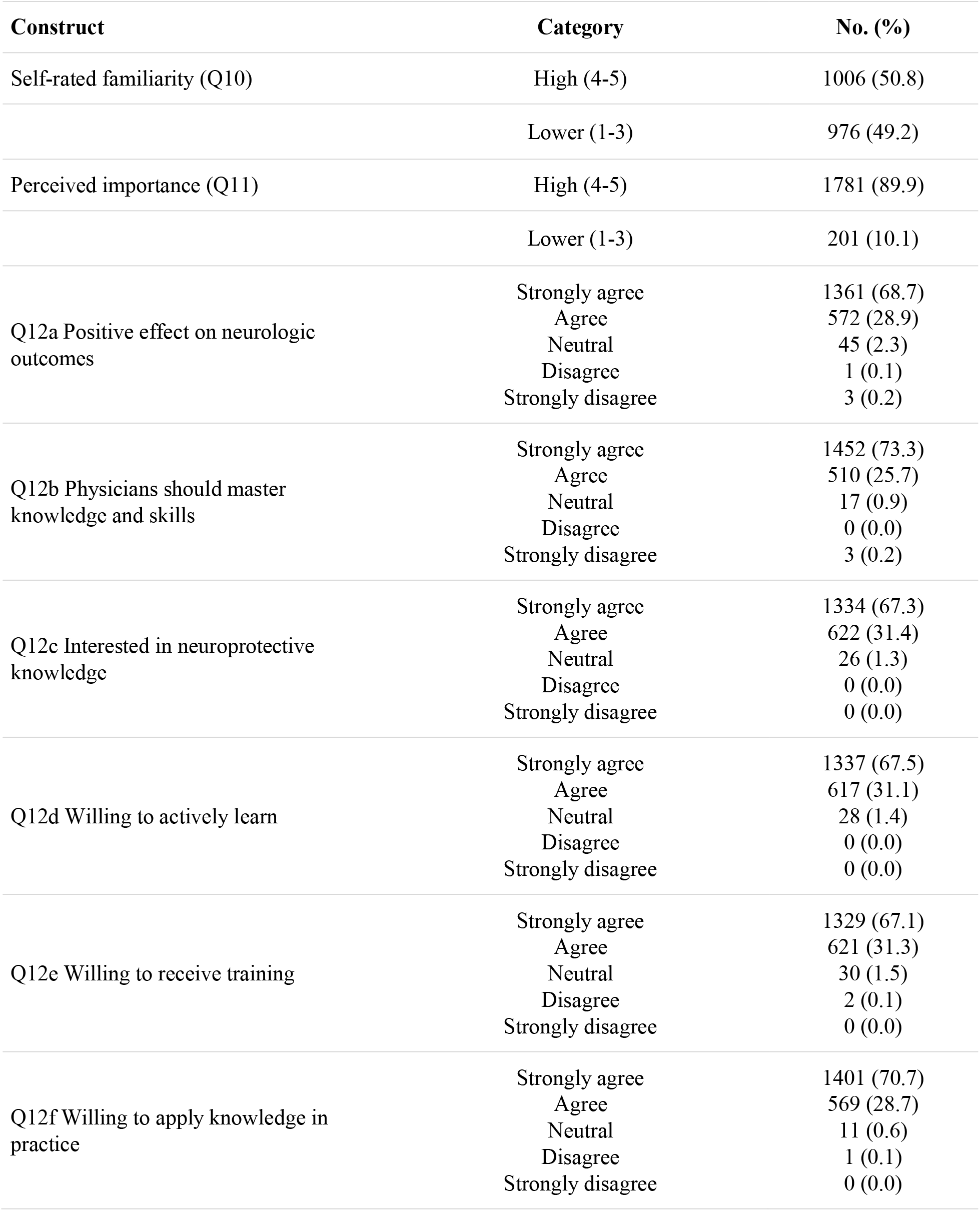
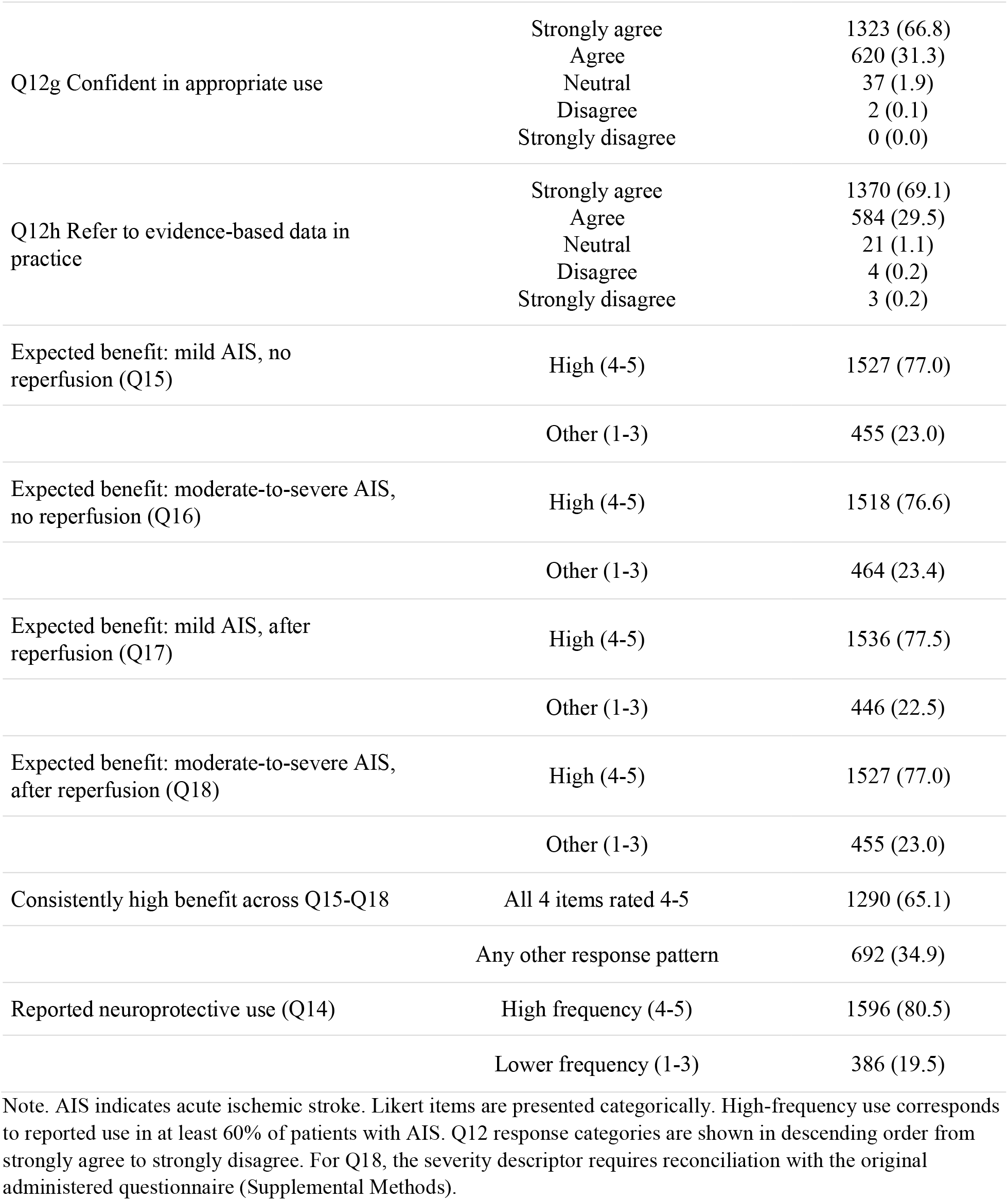
Familiarity, Perceived Importance, Expected Benefit, and Reported Prescribing (N = 1982)

High expected benefit was reported by 1527 respondents (77.0%) for mild AIS without reperfusion, 1518 (76.6%) for moderate-to-severe AIS without reperfusion, 1536 (77.5%) for mild AIS after reperfusion, and 1527 (77.0%) for moderate-to-severe AIS after reperfusion. Consistently high expected benefit across all 4 scenarios was reported by 1290 respondents (65.1%).

High-frequency neuroprotective use was reported by 1596 respondents (80.5%). In the familiarity-practice matrix, 853 (43.0%) reported high familiarity and high use, 743 (37.5%) reported low familiarity and high use, 153 (7.7%) reported high familiarity and lower use, and 233 (11.8%) reported low familiarity and lower use (Figure 1). The difference between the 2 discordant cells was 590 respondents, or 29.8 percentage points of the total sample. This contrast was descriptive.

**Figure 1.**
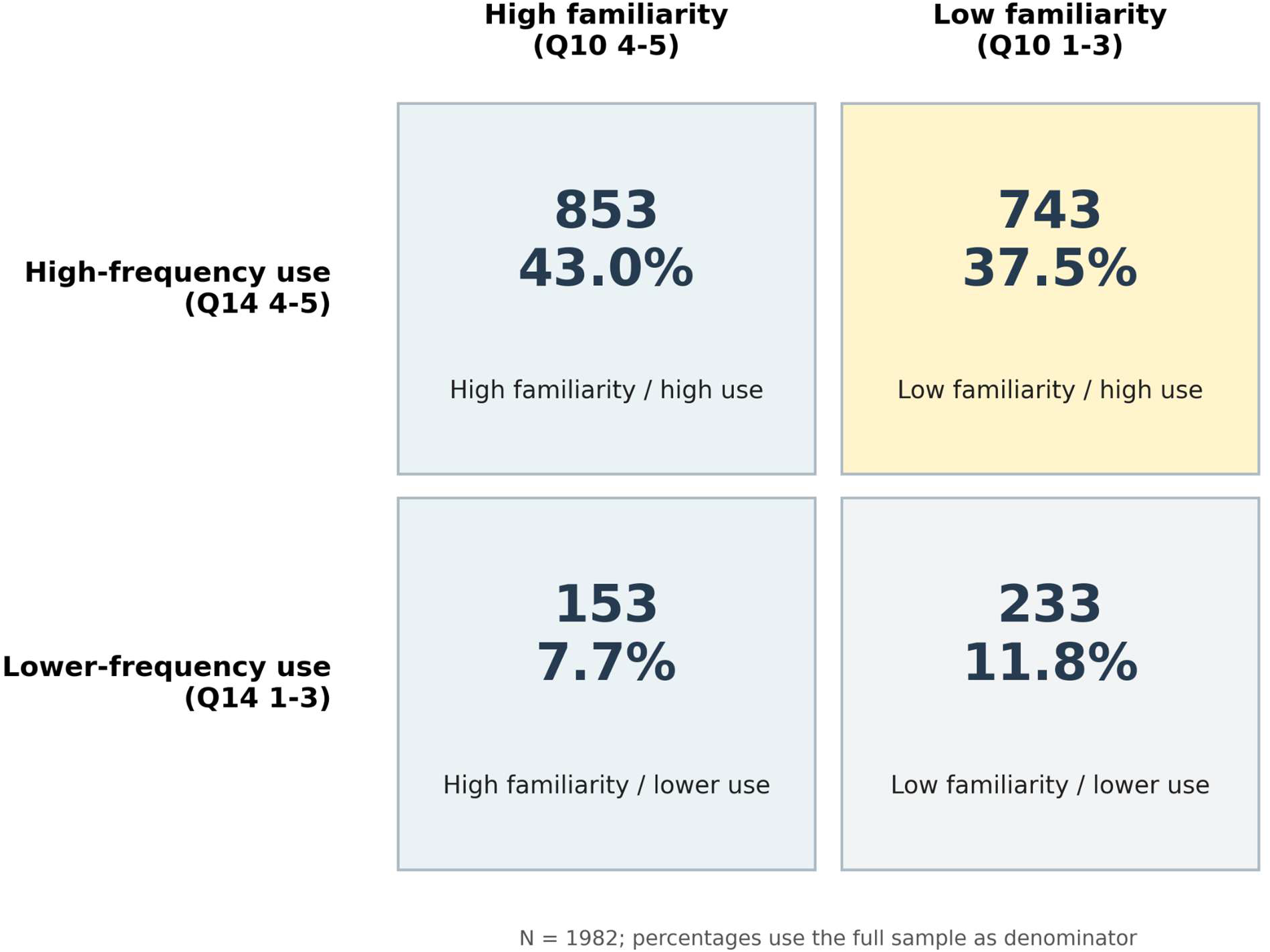
Self-rated familiarity and reported neuroprotective prescribing frequency. High familiarity was defined as Q10 ratings of 4 or 5 and lower familiarity as ratings of 1 to 3. High-frequency use was defined as Q14 ratings of 4 or 5, corresponding to use in at least 60% of patients with acute ischemic stroke; lower-frequency use comprised ratings of 1 to 3. Percentages use all 1982 respondents as the denominator. The difference between discordant cells is descriptive.

The Spearman correlation between ordinal familiarity (Q10) and prescribing frequency (Q14) was 0.144 (hospital-cluster bootstrap 95% CI, 0.099-0.188). Correlations between Q14 and the scenario-specific expected-benefit items were 0.308 (95% CI, 0.265-0.350) for Q15, 0.283 (95% CI, 0.238-0.326) for Q16, 0.289 (95% CI, 0.245-0.330) for Q17, and 0.285 (95% CI, 0.239-0.328) for Q18.

### Cluster-Adjusted Associations

In the modified Poisson GEE model, high self-rated familiarity was associated with high-frequency use (adjusted PR, 1.053; 95% CI, 1.007-1.102; P = .024), as were high perceived importance (adjusted PR, 1.294; 95% CI, 1.148-1.458; P < .001) and consistently high expected benefit (adjusted PR, 1.198; 95% CI, 1.131-1.269; P < .001) (Table 3). Compared with respondents younger than 40 years, those aged 40 to 49 years (adjusted PR, 0.921; 95% CI,0.870-0.975; P = .004) and 50 years or older (adjusted PR, 0.838; 95% CI, 0.754-0.932; P = .001) had a lower prevalence of high-frequency use; other covariates were not statistically distinguishable from their reference groups. The principal exposures retained positive associations in the sensitivity analyses. With Q14 = 5 (>90% of patients) as the outcome, the adjusted PRs were 1.190 (95% CI, 1.067-1.327) for high self-rated familiarity, 1.332 (95% CI, 1.039-1.708) for high perceived importance, and 1.719 (95% CI, 1.464-2.018) for consistently high expected benefit. With Q14 ≥3 (≥30% of patients) as the outcome, the corresponding adjusted PRs were 1.017 (95% CI, 1.004-1.030), 1.053 (95% CI, 1.006-1.102), and 1.041 (95% CI, 1.023-1.060), respectively. Replacing the consistently high expected-benefit variable with each scenario-specific high-benefit item yielded adjusted PRs for the benefit variable ranging from 1.172 (95% CI, 1.097-1.252) to 1.260 (95% CI, 1.168-1.360). Results were also similar when an independent working correlation structure was used (Table S1). Fitted means exceeded 1 for 522 records in the ≥30% outcome model and 49 in the working-independence model, which limits their interpretation; none exceeded 1 in the primary model (Table S13). The additional Q18-omission and repeated-entry checks gave similar estimates (Table S1).

**Table 3.** Adjusted Prevalence Ratios for High-Frequency Neuroprotective Use (N = 1982)

| **Variable** | **Adjusted PR (95% CI)** | **P value** |
|---|---|---|
| High self-rated familiarity (Q10 4-5 vs 1-3) | 1.053 (1.007-1.102) | .024 |
| High perceived importance (Q11 4-5 vs 1-3) | 1.294 (1.148-1.458) | <.001 |
| Consistently high expected benefit (all Q15-Q18 4-5 vs other) | 1.198 (1.131-1.269) | <.001 |
| Age 40-49 y vs <40 y | 0.921 (0.870-0.975) | .004 |
| Age ≥50 y vs <40 y | 0.838 (0.754-0.932) | .001 |
| Female vs male | 1.002 (0.962-1.043) | .925 |
| Associate chief vs attending neurologist | 0.985 (0.931-1.043) | .606 |
| Chief vs attending neurologist | 0.967 (0.892-1.048) | .407 |
| Tertiary vs secondary hospital | 1.001 (0.926-1.083) | .976 |
| Central vs eastern China | 1.032 (0.970-1.098) | .323 |
| Western vs eastern China | 0.955 (0.901-1.012) | .118 |
| Northeastern vs eastern China | 0.932 (0.829-1.048) | .237 |

Note. The outcome is high-frequency neuroprotective use (Q14 4-5 vs 1-3). Estimates are from modified Poisson regression with a log link in a generalized estimating equation, exchangeable working correlation, hospital-level clustering (251 hospitals), and robust sandwich standard errors. The model mutually adjusted for all variables shown. All 1982 respondents were included because modeled variables had no missing data. PR indicates prevalence ratio.

### Practice and Implementation Preferences

Preferred initiation windows included 2 hours or less for 974 respondents (49.1%) and 2 to 6 hours for 884 (44.6%); multiple responses were permitted. An in-hospital course of 8 to 14 days was selected by 1066 respondents (53.8%), and 1303 (65.7%) rated postdischarge sequential therapy as somewhat or very necessary. Motor function (1875 [94.6%]) and language (1682 [84.9%]) were the most frequently selected domains perceived to improve.

Commonly selected barriers were insurance restrictions (1584 [79.9%]) and high out-of-pocket burden (1466 [74.0%]); safety in vulnerable populations was selected as a concern by 1370 (69.1%). Standardized diagnostic and treatment pathways were endorsed by 1803 respondents (91.0%), and high-quality academic conferences by 1821 (91.9%). These items measured stated preferences rather than the effectiveness of implementation strategies. Complete descriptive response distributions, including additional treatment preferences and barriers, are provided in Tables S4–S14.

## Discussion

In this cross-sectional survey of 1982 neurologists from 251 hospitals across all 31 province-level regions of mainland China, 1596 respondents (80.5%) reported prescribing neuroprotective therapy to at least 60% of patients with AIS. Although 1006 respondents (50.8%) reported high familiarity with neuroprotective therapy, 743 (37.5%) were classified in the low-familiarity/high-use group. High expected benefit was reported by 76.6% to 77.5% of respondents across each of the 4 clinical scenarios, and 65.1% rated benefit as high in all 4 scenarios. In the cluster-adjusted analysis, high self-rated familiarity, high perceived importance, and consistently high expected benefit were associated with high-frequency use. These are cross-sectional relationships and should not be interpreted as determinants of prescribing.

The low-familiarity/high-use pattern does not establish inadequate knowledge or inappropriate prescribing. Familiarity was self-assessed rather than measured using an objective knowledge assessment, and prescribing frequency was reported by clinicians rather than obtained from pharmacy or medical-record data. Furthermore, the survey did not identify the specific drug, indication, dose, treatment duration, contraindications, or supporting evidence associated with each reported prescription. The discordance could reflect institutional pathways, formulary availability, reimbursement policies, established practice routines, patient expectations, or differences in how respondents interpreted the broad term neuroprotective therapy. These explanations were not measured and cannot be distinguished using the present data.

The evidence for neuroprotective therapy is heterogeneous rather than uniform across the therapeutic class. Chinese guidelines and recent randomized clinical trials address selected agents or formulations administered to defined populations within specified treatment windows.^5-8^ Consequently, evidence supporting a particular regimen cannot be generalized to every intervention respondents may have considered neuroprotective. Similarly, the narrow range of high-benefit ratings across the 4 scenarios—76.6% to 77.5%—may be compatible with broadly held benefit expectations, but it could also reflect the broad vignette wording, limited discrimination of the response scale, or a ceiling effect. Without prespecified within-participant comparisons designed to assess scenario discrimination, these data do not establish that respondents disregarded stroke severity or reperfusion status.

The findings support evaluating strategies that calibrate prescribing to agent-, patient-, and scenario-specific evidence rather than applying a uniform effort to increase or decrease neuroprotective use. Candidate approaches include concise evidence summaries, indication-linked order prompts, standardized clinical pathways, formulary review, and audit and feedback. Although 91.0% of respondents endorsed standardized pathways and 91.9% endorsed academic conferences, these preferences indicate acceptability and do not demonstrate that either strategy changes prescribing. A cluster-randomized trial in Chinese hospitals found that a multifaceted quality-improvement intervention produced a small improvement in a composite measure of adherence to evidence-based stroke care, providing a rationale for prospectively testing system-level interventions in this setting.^14^ A 2021 Cochrane review similarly concluded that educational meetings probably produce small, variable improvements in professional practice.^15^ Interventions targeting neuroprotective prescribing should therefore be evaluated using observed prescribing, evidence concordance, treatment costs, adverse events, and patient outcomes.

Future research should link physician-reported beliefs with patient-level pharmacy or electronic health-record data; distinguish individual drugs, indications, doses, and treatment durations; and characterize hospital-level factors such as formularies, pathways, and reimbursement policies. Longitudinal or experimental studies are needed to determine whether benefit expectations precede changes in prescribing and whether evidence-aligned interventions improve clinical outcomes. The present survey identifies hypotheses and potentially actionable targets but cannot establish prescribing mechanisms or intervention effects.

### Limitations

This study has several limitations. First, its cross-sectional design does not establish temporal sequence or causality. The adjusted prevalence ratios should therefore be interpreted as measures of association, not as evidence that familiarity or benefit expectations caused prescribing behavior.

Second, prescribing frequency, familiarity, and benefit expectations were self-reported and may be affected by recall, social desirability, and common-method bias. Reported prescribing was not validated against pharmacy, order-entry, or medical-record data. Moreover, neuroprotective therapy was assessed as a broad category without recording the specific agent, indication, dose, treatment duration, contraindications, or supporting evidence for individual prescriptions.

Consequently, the study cannot determine whether reported use was evidence-concordant, inappropriate, beneficial, or harmful.

Third, self-rated familiarity is not an objective measure of evidence knowledge. Several belief items also had responses concentrated in the higher categories, which may have limited discrimination between respondents. Although the analyses treat Likert responses ordinally or categorically, categorization reduces information and makes estimates potentially sensitive to the selected thresholds. Alternative cut points and scenario-specific analyses should therefore be treated as sensitivity analyses. Favorable wording in some response anchors and unresolved scenario wording may also have influenced ratings.

Fourth, the hospital-based quota recruitment strategy was designed to provide geographic and institutional coverage rather than a probability sample representative of the national neurologist workforce. Because no complete list of individually invited neurologists was available, an individual-level response rate could not be calculated, characteristics of nonrespondents could not be compared, and the direction of potential selection bias cannot be determined. In addition, 1745 respondents (88.0%) practiced in tertiary hospitals. The findings therefore may not generalize to neurologists in primary hospitals, underrepresented secondary or rural hospitals, physicians below attending level, or health systems with different formularies and reimbursement arrangements.

Fifth, residual confounding remains possible despite adjustment for physician demographic characteristics, professional title, hospital level, and geographic region. Potentially relevant factors—including hospital formularies, institutional pathways, local prescribing policies, patient case mix, workload, drug availability, reimbursement, and out-of-pocket costs—were not incorporated into the primary model. The GEE analysis accounts for statistical correlation among physicians within the same hospital but does not address unmeasured confounding or selection bias. Finally, the survey collected no patient-level treatment or outcome data; therefore, it cannot determine the effectiveness, safety, cost-effectiveness, or clinical consequences of the reported prescribing patterns.

## Data Availability

The datasets generated and/or analyzed during the current study are available from the corresponding author on reasonable request.

## Non-standard Abbreviations and Acronyms

AIS: acute ischemic stroke
GEE: generalized estimating equations
PR: prevalence ratio
STROBE: Strengthening the Reporting of Observational Studies in Epidemiology

## Acknowledgments

The Chinese Stroke Association coordinated survey distribution.

